# A Cost of Illness Analysis of Children with Encephalitis Presenting to A Major Hospital in Vietnam

**DOI:** 10.1101/2024.04.24.24306275

**Authors:** Nguyen Hoang Thien Huong, Nguyen Duc Toan, Truong Huu Khanh, Le Quoc Thinh, Le Nguyen Thanh Nhan, Ngo Ngoc Quang Minh, Nguyen Thi Kim Thoa, Nguyen Thanh Hung, Du Tuan Quy, C. Louise Thwaites, Sarosh R Irani, Le Van Tan, Hugo C. Turner

**Affiliations:** Oxford University Clinical Research Unit, Ho Chi Minh City, Vietnam; Children’s Hospital 1, Ho Chi Minh City, Vietnam; Department of Paediatrics, Vietnam National University Ho Chi Minh City School of Medicine, Ho Chi Minh City, Vietnam; Department of Paediatrics, Pham Ngoc Thach University of Medicine, Ho Chi Minh City, Vietnam; Nuffield Department of Medicine, University of Oxford, Oxford, UK; Departments of Neurology and Neurosciences, Mayo Clinic, Jacksonville, Florida, US; MRC Centre for Global Infectious Disease Analysis, School of Public Health, Imperial College London, London, UK

**Keywords:** Encephalitis, Children, Economic Burden, Cost of Illness, Vietnam

## Abstract

**Background:** Encephalitis is a significant global health problem, especially in children. Knowledge of its economic burden is essential for policymakers in prioritizing the development and implementation of interventions but remains limited.

**Methods:** An observational study was prospectively conducted at a major children’s hospital in Ho Chi Minh City, Vietnam, from 2020 to 2022. Data on direct medical costs, direct non-medical costs and productivity costs were collected alongside demographic information, clinical features, diagnosis, severity, and outcomes of study participants. This was used to undertake a cost of illness analysis from a societal perspective.

**Results:** Data were collected from a total of 164 paediatric patients. The mean cost of illness per case was estimated at US$2,820.43 (95% confidence interval (CI), US$2,431.96– US$3,208.91), of which productivity costs accounted for US$434.04 (95% CI, US$362.48– US$505.60). The direct costs were the main cost driver, accounting for 84.6% of the total cost of illness (US$2,386.38 (95%CI: US$2,033.91–US$2,738.85)). The cost of illness was higher in more severe patients, patients with sequelae, and ventilated patients. On average 51.8% of direct medical costs attributed to hospitalisation (US$960.09) resulted in out-of-pocket payments from the patient’s family.

**Conclusions:** The results showed that the cost of illness of encephalitis in children is considerable. The results will be useful for policymakers in prioritizing resources for the development and implementation of intervention strategies to reduce the burden of paediatric encephalitis.

## INTRODUCTION

Encephalitis is responsible for a significant global burden of disease, especially in children (1). An estimated 4 to 10 children per 100,000 are hospitalized with encephalitis every year (2–4) and it is associated with mortality rates of 2 to 31% (5–7). In a retrospective analysis of encephalitis cases in England and Wales in children, the annual mortality rate was 0.07 (95% CI: 0.05–0.08) per 100,000 population aged between 0–17 years (8). Amongst 7,298 children in the United States admitted with encephalitis between 2004 and 2013, 2,933 patients (40%) required treatment in a paediatric intensive care unit, and had an overall median length of hospital stay of 16 days (4). This high requirement for specialised treatment and lengthy hospital stay is linked to the high cost of encephalitis treatment (2,4,9–11).

Data from the United States, based on a nationally representative database of hospitalizations the total cost of encephalitis-associated hospitalizations was estimated to be US$2.0 billion in 2010 (12) and the average economic burden of a child with encephalitis was estimated to be between US$64,000 and US$260,000 depending on the level of healthcare and rehabilitation (2,4,10). In low- and middle-income countries (LMICs) where encephalitis in children is common, there are only limited data available (13–15). A longitudinal follow-up study in Nepal of acute encephalitis syndrome in children from two hospitals in Nepal (13) where the median out-of-pocket cost to families, including medical bills, medication and lost earnings was US$1,151 for children with severe/moderate impairment and US$524 for those with mild/no impairment (13). In a Cambodian study that interviewed affected families, it was found that costs related to acute hospital admission were the major cost driver (14). Medication was identified to be the predominant acute medical cost associated with paediatric encephalitis in Indonesia (15).

A previous cost of illness analysis of Japanese encephalitis in Vietnam estimated a mean total cost of US$3,371 per acute episode (16). This showed that Japanese encephalitis patients and families in Vietnam suffer notable medical, economic, and social hardship (16). Whilst this study has provided valuable insight into the costs of one cause of encephalitis in Vietnamese children, knowledge regarding the economic burden of paediatric encephalitis more generally is lacking, but essential to inform local policymakers in planning and prioritizing resources for diagnosis and treatment and public health interventions. For this reason, we carried out this study, conducting a prospective observational study to estimate the costs attributed to paediatric encephalitis in a major hospital in Southern Vietnam.

## METHODS

### Study design and setting

The study was a prospective observational study, conducted at Children’s Hospital 1 (CH1) in Ho Chi Minh City in Vietnam between January 2020 and December 2022 (17). CH1 is a 1,600- bed hospital, and is the largest tertiary hospital for children living in the southern provinces of Vietnam, with a catchment population of over 40 million. Annually, CH1 has approximately 90,000 admissions. Of these, approximately 150–200 cases receive a diagnosis of encephalitis.

### Consent

Written informed consent will be obtained from all study participants or their representatives before any data from patients is collected for the study. The study staff will discuss the study with the parent/representative of potential participants. Study staff will describe the purpose of the study, the study procedures, possible risks/benefits, the rights and responsibilities of patient, and alternatives to enrolment. The parent/representative will be invited to ask questions which will be answered by study staff, and they will be provided with appropriate numbers to contact if they have any questions subsequently. If the parent/representative agrees for their child to participate, they will be asked to sign and date two copies of an informed consent form. A copy of the form will be given to them to keep. If required, the parent/representative will be given up to 48 hours to consider for their children to take part in the study. In addition to the procedures above, illiterate signatories will have the informed consent form read to them in the presence of a witness who will sign to confirm that the form was read accurately and that the participant or representative agrees to participation. All informed consent forms will be written in the local language and will use terms that are easily understandable.

### Inclusion and exclusion criteria

Any child admitted to the Department of Infectious Diseases and Neurology of Children’s Hospital 1 and fulfilling the case definition of encephalitis between January 2020 to December 2022 (Supplemental Figure 1) (17–19) was eligible for study participation.

Patients were excluded if no informed consent was obtained. Given the descriptive nature of the present study, we used a convenience sampling approach, and no formal sample size calculation was performed.

### Data collection

Clinical data including dates of birth, admission and discharge, demographic data, clinical features, routine diagnosis, and outcomes were collected from all participants. The definitions of clinical outcomes in our study can be found in Supplemental Table S1. Patient consciousness level was evaluated using the Paediatric Glasgow coma scale (GCS) and those with GCS < 9 were categorised as having severe disease (20). The modified Rankin scale (mRS) for children was used to assess the degree of disability or dependence in daily activities, and scores ≥ 3 were categorised as severe disability (21). Requiring respiratory support with mechanical ventilation was used as a proxy indicator of disease severity (9,22–24). Data were collected from hospital invoices and structured questionnaires administered through face-to-face interviews. Caregivers were interviewed at admission (for the costs before hospital admission), just prior to hospital discharge (for the costs during hospitalization), and at the outpatient clinic of the hospital (for the costs up to seven days after discharge). We did not carry out telephone interviews to follow up on these patients.

### Laboratory diagnosis

As part of routine care at CH1, encephalitis diagnosis was made through analysis of cerebrospinal fluid (CSF) including culture and/or microscopy for detection of bacterial infections. Patients with suspected encephalitis were further tested for herpes simplex virus and Japanese encephalitis virus using PCR and serology, respectively. Patients with clinical presentations compatible with autoimmune encephalitis were tested for possible forms of the disease using commercial fixed cells-based assays (EUROIMMUN, Lübeck, Germany) (25). Auto-antibody testing was conducted on the cerebrospinal fluid (CSF) samples only.

### Cost evaluation

An overview of the cost components considered is presented in Figure 1. The total cost of illness consisted of direct medical costs, direct non-medical costs and productivity costs (Figure 1). The costs were collected from a societal perspective. Data on the cost of illness was collected capturing the period prior to hospital admission, during hospitalization, and seven days after discharge.

**Figure 1.**
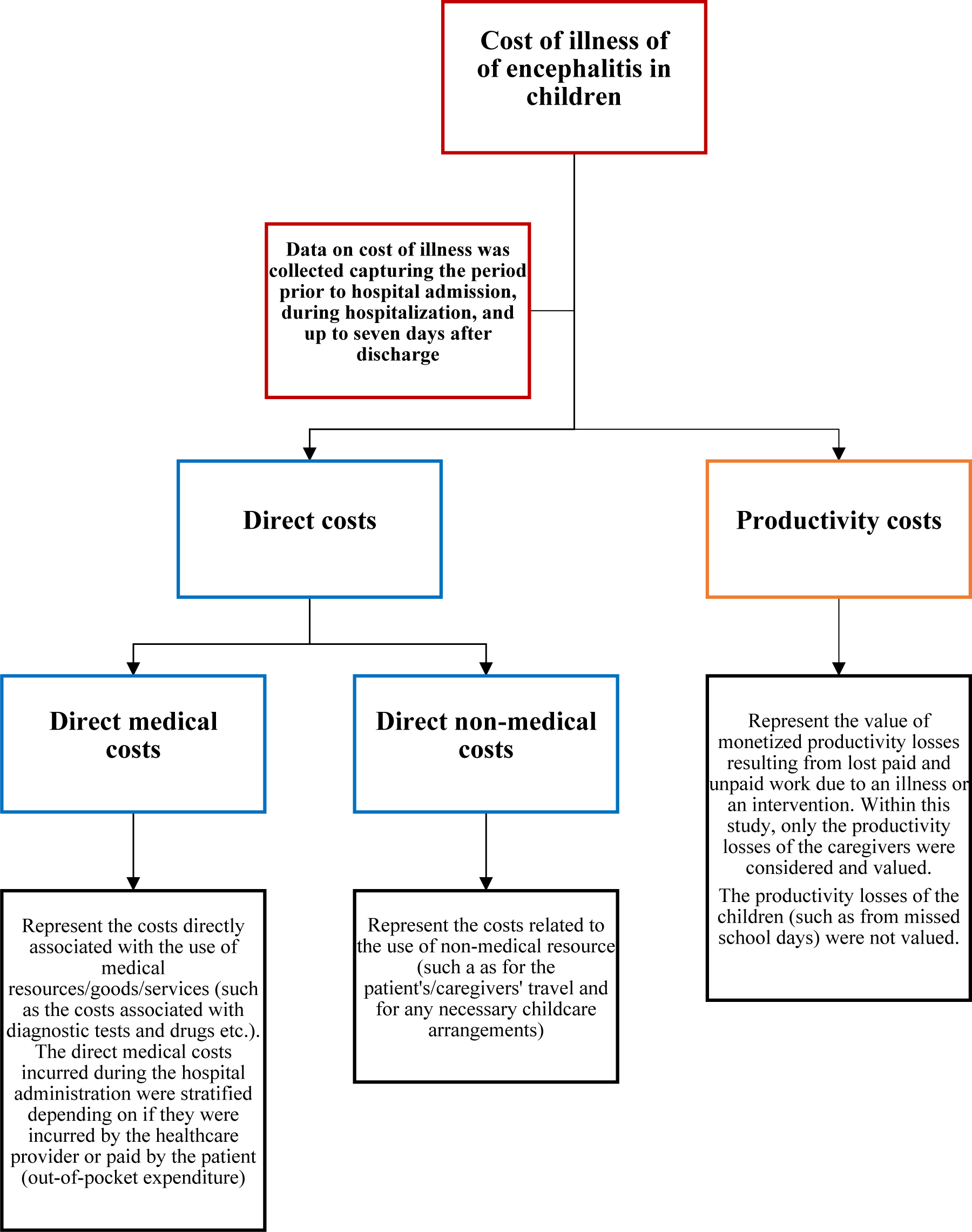
An overview of the components considered within the cost of illness calculation.

The direct medical costs consist of the costs directly associated with the use of medical resources/goods/services (such as the costs associated with diagnostic tests and drugs etc.). These costs were calculated based on the hospital invoices and the information obtained from the face-to-face interviews (Figure 1). The direct medical costs attributed to hospitalisation were collected based on the official invoices within the patients’ in-hospital medical records. These costs could be stratified by the cost covered by the government’s health insurance programme and the family’s out-of-pocket payments. The direct medical costs incurred before hospital admission and up to seven days after discharge were captured through interviews. However, because these were not based on official invoices it was not possible to stratify these costs depending on if they were paid by the families themselves vs covered by the health insurance programme.

The direct non-medical costs represent the costs related to the use of non-medical resources. These include the patients’/caregivers’ travel costs and other expenses related to the care of the patient. Data on direct non-medical costs was collected through face-to-face interviews (Figure 1).

The productivity costs represent the value of monetized productivity losses resulting from lost paid and unpaid work due to an illness or an intervention. Within this study, only the productivity losses of the caregivers were considered and valued. The productivity losses of children (such as from missed school days) were not valued. Data on the caregivers’ productivity losses incurred was collected through face-to-face interviews by asking about the number of days they lost due to caregiving before hospital admission, during hospitalization, and up to seven days after discharge. How these productivity losses were valued depended on the activity the informal caregivers reported giving up. For those who reported giving up paid employment, their losses were valued based on their reported monthly salary. For those who reported losing unpaid work, their losses were valued based on the minimum wage. As the minimum wage in Vietnam varies across the different provinces (Supplemental Table S2) (26), we used the monthly minimum wage that corresponded to the address (province) of the patient. The monthly salary and minimum wage values were adjusted to a daily value based on the caregivers’ reported number of working days per month (Supplemental Table S3). The productivity costs were calculated based on this daily value multiplied by the reported number of days lost.

Due to the local COVID-19 regulations during the study, only one caregiver (usually the father or mother) was allowed to stay with and take care of the child and the caregiver did not change during the hospitalisation period. No excess in-hospital death was considered in the calculation of productivity costs.

Under the government health insurance scheme in Vietnam, children under 6 years old are provided with free health care services. For children from 6 to 14 years old, there are two main health insurance programs, which are operated by the Vietnam Health Insurance Organization (VHI) on a non-profit and public basis. However, the families have to pay for medical services that are not listed in the health insurance directory issued by the Ministry of Health (27).

We summarized all values of illness costs as means and 95% confidence intervals (CIs) in US dollars (US$) with a conversion rate of US$1 equivalent to 23,271.2 dong (the exchange rate between Vietnamese Dong and US dollars for the year 2022 based on the database from the World Bank. Costs incurred in 2020 and 2021 were adjusted to 2022 prices using GDP deflators for Vietnam using the approach outlined within Turner et al (28).

### Ethical Statement

The Institutional Review Board of Children’s Hospital 1 (1503/QD-BVND1) and the Oxford Tropical Research Ethics Committee (OxTREC 7-20) approved the study. The collection of all biological samples was performed as part of routine clinical assessments and was consistent with the local standard of care and good clinical practice. Written informed consent was obtained from all study participants or their representatives before any data from patients was collected for the study.

## RESULTS

### Baseline features of children with encephalitis

Between January 2020 and December 2022, 164 children with clinical features of encephalitis were enrolled in the study. Among these, 23/164 patients (14.0%) were confirmed as NMDAR-antibody encephalitis. Viral encephalitis was diagnosed in 18/164 cases (11.0%). The other cases were reported as unknown infectious aetiology encephalitis (123/164, 75%) as the aetiologies of these cases could not be identified (Table 1). The baseline features of children with encephalitis are shown in Table 1. The residences of children with encephalitis and the distance to our children’s hospital can be found in Supplemental Figure S2.

**Table 1.** Baseline features of the sample.

|  | **NMDAR-antibody**<br>**encephalitis**<br>**(n = 23)** | **Confirmed viral**<br>**encephalitis**<br>**(n = 18)** | **Unknown aetiology**<br>**encephalitis**<br>**(n = 123)** |
| --- | --- | --- | --- |
| **General features** |  |  |  |
| **Gender** |  |  |  |
| Male<sup>a</sup> | 4 (17.4) | 13 (72.2) | 68 (55.3) |
| Female<sup>a</sup> | 19 (82.6) | 5 (27.8) | 55 (44.7) |
| **Age (years)<sup>b</sup>** | 9.8 (7.8–11.7) | 8.4 (6.4–10.5) | 8.6 (7.8–9.4) |
| **Residence** |  |  |  |
| Ho Chi Minh city<sup>a</sup> | 7 (30.4) | 0 (0.0) | 36 (29.3) |
| Other provinces<sup>a</sup> | 16 (69.6) | 18 (100.0) | 87 (70.7) |
| **Clinical features** |  |  |  |
| **Glasgow coma scale**<br>**(GCS)<sup>b</sup>** | 11 (10–12) | 8 (7–10) | 12 (11– 2) |
| GCS <9<sup>a</sup><br>(more severe) | 4 (17.4) | 11 (61.1) | 11 (8.9) |
| GCS ≥9<sup>a</sup><br>(less severe) | 19 (82.6) | 7 (38.9) | 112 (91.1) |
| Ventilated<sup>a</sup> | 6 (26.1) | 11 (61.1) | 14 (11.4) |
| Not ventilated<sup>a</sup> | 17 (73.9) | 7 (38.9) | 109 (88.6) |
| **Outcomes** |  |  |  |
| **Modified Rankin scale**<br>**(mRS)<sup>b</sup>** | 2 (1–2) | 1 (1–2) | 1 (0–1) |
| mRS <3<sup>a</sup><br>(less severe) | 16 (69.6) | 16 (88.9) | 109 (88.6) |
| mRS ≥3<sup>a</sup><br>(more severe) | 7 (30.4) | 2 (11.1) | 14 (11.4) |
| **Sequelae<sup>a</sup>** | 13 (56.5) | 12 (66.7) | 39 (31.7) |
| **Complications<sup>a</sup>** | 18 (78.3) | 12 (66.7) | 66 (53.7) |
| **In hospital death<sup>a</sup>** | 1 (4.3) | 0 (0.0) | 2 (1.6) |
| **Duration of hospital**<br>**stay (days)<sup>b</sup>** | 37 (27–47) | 23 (13–33) | 19 (16–22) |

<sup>a</sup>output presented as n (%)
<sup>b</sup>output being presented as mean (95% CI)
Abbreviation: CI, confidence interval

### The informal caregivers’ characteristics

The most common normal activities the caregivers reported giving up to provide care for the child were paid employment (76/164, 46.3%), housework (36/164, 22.0%) and subsistence farming (32/164, 19.5%). The mean number of days lost was 36.7 days (95% CI: 32.6–40.7 days). The productivity losses were stratified depending on whether they were incurred before hospitalization, during hospitalization, or after hospitalization. The mean monthly income of the caregivers was US$276.35 (95% CI: US$251.09–US$301.60) (Table 2).

**Table 2.** Information regarding the caregivers.

| **Characteristics (N = 164\*)** | **n (%) or mean (95% CI)** |
| --- | --- |
| **Residence (n, (%))** |  |
| Ho Chi Minh City | 43 (26.2) |
| Other provinces | 121 (73.8) |
| **Relationship (n, (%))** |  |
| Mother | 118 (72.0) |
| Father | 46 (28.0) |
| **Normal activities (n, (%))** |  |
| Paid employment | 77 (46.9) |
| Business ownership | 19 (11.6) |
| Subsistence farming (unpaid work) | 32 (19.5) |
| Housework (unpaid work) | 36 (22.0) |
| **Working days per month (mean, (95% CI))** | 25.5 (25.1–25.8) |
| Working days per month of paid workers | 25.7 (25.3–26.2) |
| Working days per month of unpaid workers | 25.1 (24.6–25.7) |
| **Days of work lost (mean, (95% CI))** | 36.7 (32.6–40.7) |
| Days lost before hospitalization | 6.9 (5.7–8.0) |
| Days lost during hospitalization | 20.6 (17.7–23.5) |
| Days lost after hospitalization | 9.2 (7.9–10.5) |
| **Individual income per month (mean US\$ (95% CI))** | 276.35 (251.09–301.60) |

*Abbreviation: CI, confidence interval*
*\*Only one registered caregiver could stay with the child during the hospitalisation period.*

### An overview of the costs of illness attributed to paediatric encephalitis

A summary of the estimated costs associated with encephalitis in children is presented in Table 3. The total mean cost of illness for each case was estimated to be US$2,820.43 (95%CI: US$2,431.96–US$3,208.91). The direct costs constituted the majority (84.6%) of this total cost of illness with a mean of US$2,386.38 (95%CI: US$2,033.91–US$2,738.85). In terms of direct costs, the direct medical cost incurred during hospitalization was found to be the main component/driver, with a mean of US$1,853.24 (95%CI: US$1,525.33–US$2,181.15). Importantly, in Vietnam, the government’s health insurance programme does not cover all the medical costs and in this study, the patients had to pay on average 51.8% (US$960.09) of the direct medical costs incurred during hospitalization. The mean direct non-medical cost was US$398.56 (95%CI US$359.62–US$437.50) per case – 44.1% of which were related to transportation. The remaining accounted for other types of direct non-medical costs – such as costs related to the care of the patient. The productivity costs during hospitalization were the dominant contributor to the total productivity costs (US$248.54 of US$434.04).

**Table 3.** Summary of the estimated costs of illness of paediatric encephalitis cases.

| **Costs type** | **Mean US\$ (95% CI) per case** |
| --- | --- |
| **Direct medical cost** | 1,987.82 (1,661.13–2,314.51) |
| Direct medical costs before hospital admission | 84.99 (63.27–106.72) |
| Direct medical costs during hospitalization | 1,853.24 (1,525.33–2,181.15) |
| Direct medical costs up to seven days after discharge | 49.57 (34.79–64.35) |
| **Direct medical cost paid by the patients** |  |
| Direct medical costs before hospitalization paid by the patients | NA |
| Direct medical costs during hospitalization paid by the patients | 960.09 (633.34–1,286.84) |
| Direct medical costs up to seven days after discharge paid by the patients | NA |
| **Direct non-medical cost** | 398.56 (359.62–437.50) |
| Transportation costs before hospital admission | 23.97 (18.67–29.26) |
| Transportation costs during hospitalization | 110.67 (97.85–123.48) |
| Transportation costs up to seven days after discharge | 41.21 (32.95–49.47) |
| Other expenses<sup>1</sup> | 222.70 (194.32–251.09) |
| **Total productivity cost** | 434.04 (362.48–505.60) |
| Productivity costs before hospital admission | 76.81 (61.90–91.71) |
| Productivity costs during hospitalization | 248.54 (199.72–297.37) |
| Productivity costs up to seven days after discharge | 108.69 (87.10–130.27) |
| **Total direct cost** | 2,386.38 (2,033.91–2,738.85) |
| **Total costs of illness** | 2,820.43 (2,431.96–3,208.91) |

*Abbreviation: CI, confidence interval*
*NA: Not available*
*<sup>1</sup> Such as other costs related to the care of the patient.*

### The valuation of productivity costs

Paid work constituted 58.5% (96/164) of the normal activities of the caregivers. The mean productivity cost for caregivers who lost paid work was estimated to be US$563.66 based on their reported wages. For those who lost unpaid work, the mean productivity costs were calculated as US$251.06 based on the daily minimum wage. Consequently, the productivity costs of paid workers are estimated to be twice as much as caregivers performing unpaid work (Table 4). The productivity loss during hospitalization contributed to over half of the total productivity loss of caregivers (Table 4).

**Table 4.** The valuation of the caregivers’ productivity costs.

|  | **Caregivers losing paid work<sup>a</sup>** | **Caregivers losing unpaid work<sup>b</sup>** |
| --- | --- | --- |
|  | Mean (95% CI) per caregiver | Mean (95% CI) per caregiver |
| **Monthly income - US\$** | 352.05 (315.96–388.13) | 169.47 (164.12–174.83) |
| **Working days per month** | 25.7 (25.3–26.2) | 25.1 (24.6–25.7) |
| **Daily value of a lost day<sup>c</sup> - US\$** | 14.25 (12.76–15.74) | 7.01 (6.68–7.35) |
| **Days lost before hospitalisation** | 6.7 (5.4–8.0) | 7.1 (4.9–9.2) |
| **Days lost during hospitalisation** | 21.7 (17.6–25.8) | 19.1 (14.9–23.2) |
| **Days lost after hospitalisation** | 9.5 (7.6–11.4) | 8.8 (7.3–10.2) |
| **Total days lost** | 37.9 (32.3–43.5) | 35.0 (29.0–40.9) |
| **Productivity cost before hospitalisation - US\$** | 95.41 (72.83–117.99) | 50.54 (35.39–65.69) |
| **Productivity cost during hospitalisation - US\$** | 326.54 (250.15–402.92) | 138.43 (103.05–173.82) |
| **Productivity cost after hospitalisation - US\$** | 141.70 (106.96–176.44) | 62.08 (50.63–73.53) |
| **Total productivity cost - US\$** | 563.66 (452.78–674.53) | 251.06 (201.84–300.29) |

<sup>a</sup>For those that reported losing paid work, their productivity losses were valued based on the monthly salary (n=96).
<sup>b</sup>For those that reported losing unpaid work, their productivity losses were valued based on the monthly minimum wage (n=68).
<sup>c</sup>Calculated by dividing the monthly income (salary for lost paid work and minimum wage for lost unpaid work) by the reported number of working days per month (Supplemental Table S2). The productivity cost was calculated by multiplying this daily value by the number of days lost.

### Costs of illness of encephalitis in children by geographic location, diagnosis, severity, and outcomes

We identified a number of factors that influenced the projected costs of illness (Table 5). Patients with confirmed NMDAR-antibody encephalitis were associated with higher costs of illness (mean, US$4,455.19) compared to other categories of encephalitis in children. As expected, in terms of level of consciousness, the total costs of illness of more severely ill patients was higher than that of less severe patients (mean, US$4,570.56 vs mean, US$2,490.70). Ventilated patients had higher total costs of illness than that of non-ventilated patients (mean, US$4,702.71 vs mean, US$2,381.70). Assessing the costs by the degree of disability or dependence in their daily activities, the total costs of illness at discharge of more severely affected patients (mRS ≥3) were much higher than that of less severe patients (mRS <3) (mean, US$6,204.17 compared to US$2,268.47). Similarly, patients with sequelae had higher total costs compared to those of patients without sequelae (mean, US$3,642.94 vs US$2,294.02) (Table 5).

**Table 5.**
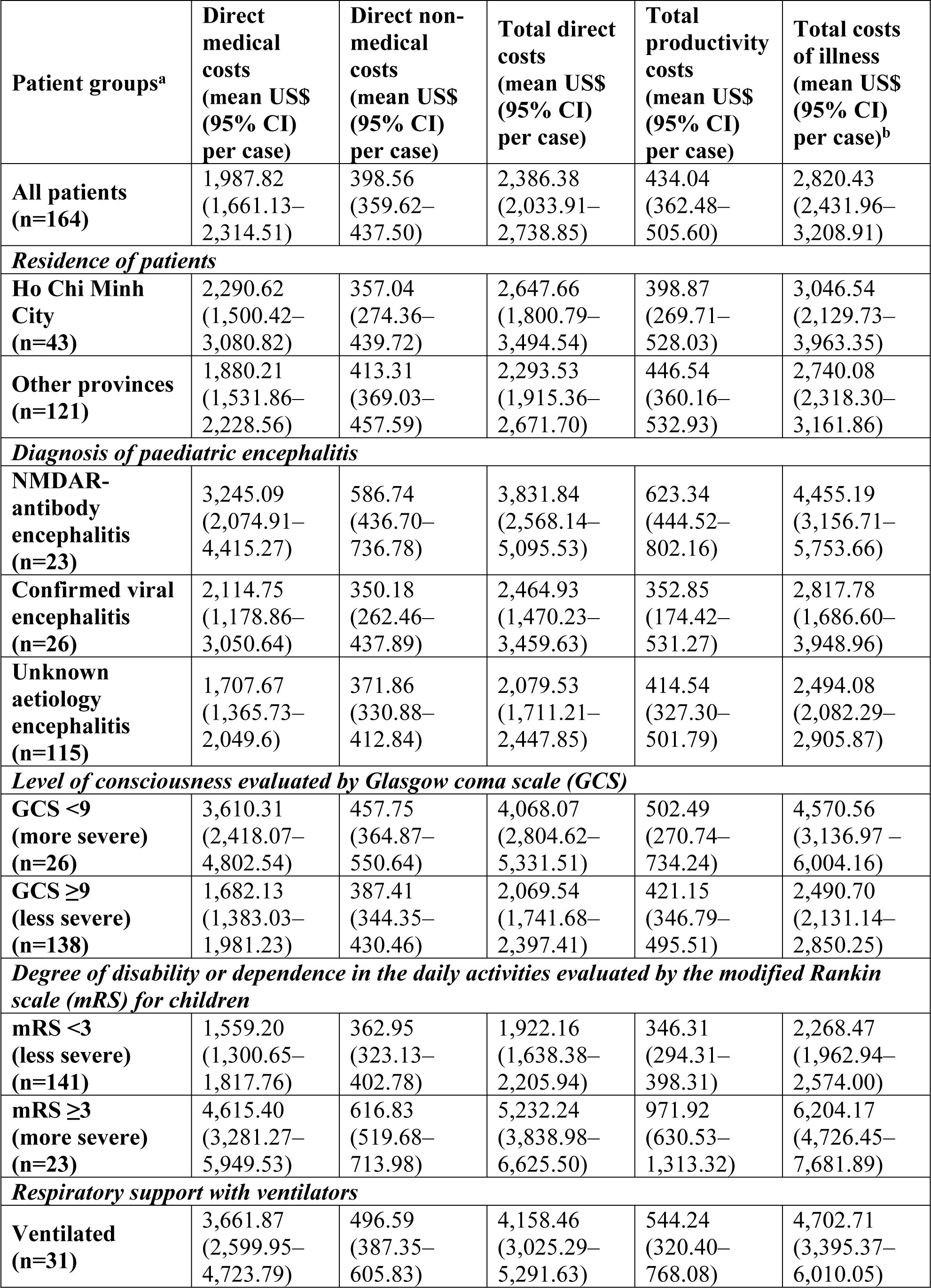
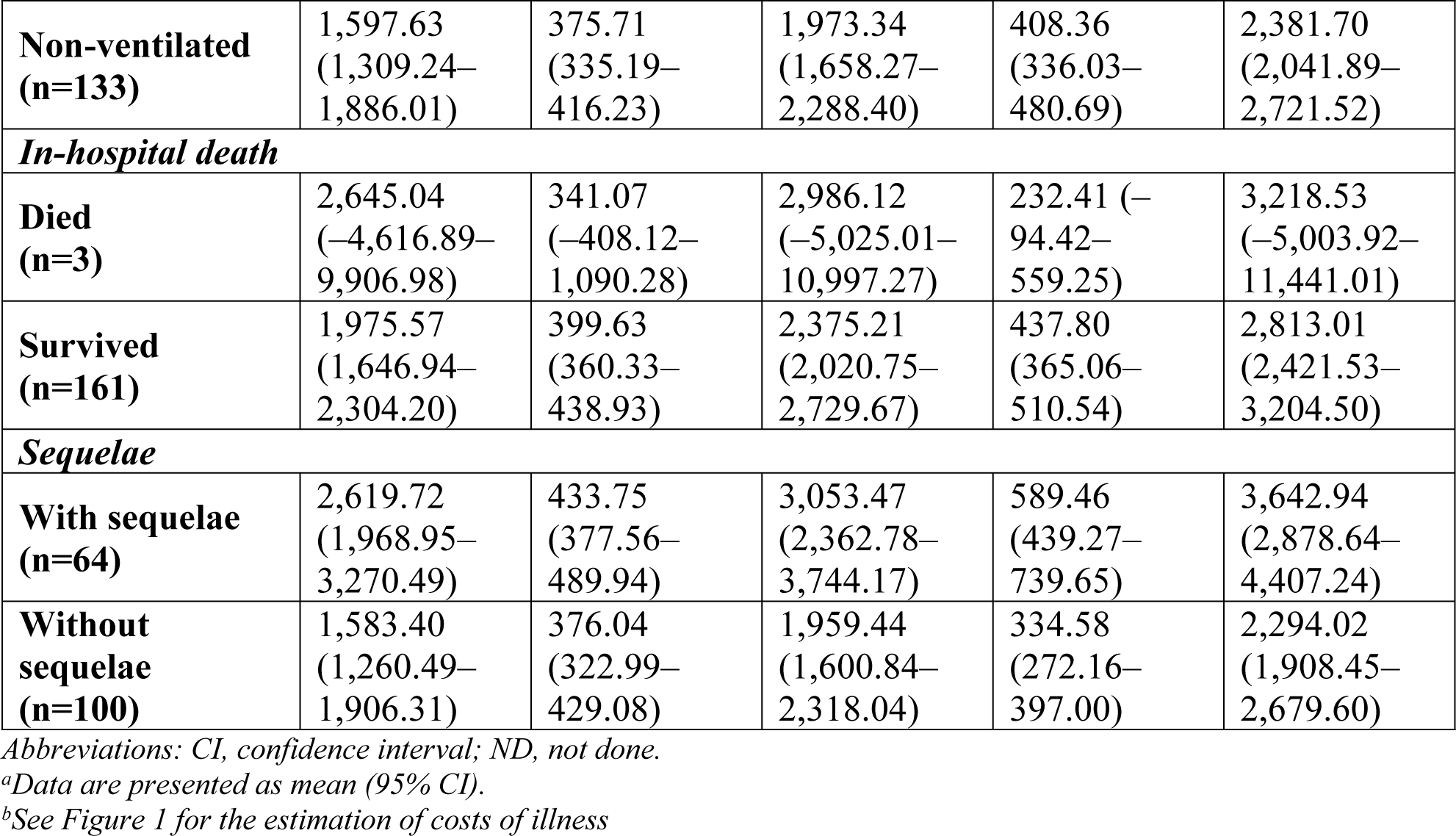
The estimated costs of illness stratified by different patient groupings.

| **Patient groups<sup>a</sup>** | **Direct medical costs (mean US\$ (95% CI) per case)** | **Direct non-medical costs (mean US\$ (95% CI) per case)** | **Total direct costs (mean US\$ (95% CI) per case)** | **Total productivity costs (mean US\$ (95% CI) per case)** | **Total costs of illness (mean US\$ (95% CI) per case)<sup>b</sup>** |
| --- | --- | --- | --- | --- | --- |
| **All patients (n=164)** | 1,987.82<br>(1,661.13–2,314.51) | 398.56<br>(359.62–437.50) | 2,386.38<br>(2,033.91–2,738.85) | 434.04<br>(362.48–505.60) | 2,820.43<br>(2,431.96–3,208.91) |
| ***Residence of patients*** |  |  |  |  |  |
| **Ho Chi Minh City (n=43)** | 2,290.62<br>(1,500.42–3,080.82) | 357.04<br>(274.36–439.72) | 2,647.66<br>(1,800.79–3,494.54) | 398.87<br>(269.71–528.03) | 3,046.54<br>(2,129.73–3,963.35) |
| **Other provinces (n=121)** | 1,880.21<br>(1,531.86–2,228.56) | 413.31<br>(369.03–457.59) | 2,293.53<br>(1,915.36–2,671.70) | 446.54<br>(360.16–532.93) | 2,740.08<br>(2,318.30–3,161.86) |
| ***Diagnosis of paediatric encephalitis*** |  |  |  |  |  |
| **NMDAR-antibody encephalitis (n=23)** | 3,245.09<br>(2,074.91–4,415.27) | 586.74<br>(436.70–736.78) | 3,831.84<br>(2,568.14–5,095.53) | 623.34<br>(444.52–802.16) | 4,455.19<br>(3,156.71–5,753.66) |
| **Confirmed viral encephalitis (n=26)** | 2,114.75<br>(1,178.86–3,050.64) | 350.18<br>(262.46–437.89) | 2,464.93<br>(1,470.23–3,459.63) | 352.85<br>(174.42–531.27) | 2,817.78<br>(1,686.60–3,948.96) |
| **Unknown aetiology encephalitis (n=115)** | 1,707.67<br>(1,365.73–2,049.6) | 371.86<br>(330.88–412.84) | 2,079.53<br>(1,711.21–2,447.85) | 414.54<br>(327.30–501.79) | 2,494.08<br>(2,082.29–2,905.87) |
| ***Level of consciousness evaluated by Glasgow coma scale (GCS)*** |  |  |  |  |  |
| **GCS <9 (more severe) (n=26)** | 3,610.31<br>(2,418.07–4,802.54) | 457.75<br>(364.87–550.64) | 4,068.07<br>(2,804.62–5,331.51) | 502.49<br>(270.74–734.24) | 4,570.56<br>(3,136.97 – 6,004.16) |
| **GCS ≥9 (less severe) (n=138)** | 1,682.13<br>(1,383.03–1,981.23) | 387.41<br>(344.35–430.46) | 2,069.54<br>(1,741.68–2,397.41) | 421.15<br>(346.79–495.51) | 2,490.70<br>(2,131.14–2,850.25) |
| ***Degree of disability or dependence in the daily activities evaluated by the modified Rankin scale (mRS) for children*** |  |  |  |  |  |
| **mRS <3 (less severe) (n=141)** | 1,559.20<br>(1,300.65–1,817.76) | 362.95<br>(323.13–402.78) | 1,922.16<br>(1,638.38–2,205.94) | 346.31<br>(294.31–398.31) | 2,268.47<br>(1,962.94–2,574.00) |
| **mRS ≥3 (more severe) (n=23)** | 4,615.40<br>(3,281.27–5,949.53) | 616.83<br>(519.68–713.98) | 5,232.24<br>(3,838.98–6,625.50) | 971.92<br>(630.53–1,313.32) | 6,204.17<br>(4,726.45–7,681.89) |
| ***Respiratory support with ventilators*** |  |  |  |  |  |
| **Ventilated (n=31)** | 3,661.87<br>(2,599.95–4,723.79) | 496.59<br>(387.35–605.83) | 4,158.46<br>(3,025.29–5,291.63) | 544.24<br>(320.40–768.08) | 4,702.71<br>(3,395.37–6,010.05) |
| **Non-ventilated**<br>**(n=133)** | 1,597.63<br>(1,309.24–<br>1,886.01) | 375.71<br>(335.19–<br>416.23) | 1,973.34<br>(1,658.27–<br>2,288.40) | 408.36<br>(336.03–<br>480.69) | 2,381.70<br>(2,041.89–<br>2,721.52) |
| ***In-hospital death*** |  |  |  |  |  |
| **Died**<br>**(n=3)** | 2,645.04<br>(–4,616.89–<br>9,906.98) | 341.07<br>(–408.12–<br>1,090.28) | 2,986.12<br>(–5,025.01–<br>10,997.27) | 232.41 (–<br>94.42–<br>559.25) | 3,218.53<br>(–5,003.92–<br>11,441.01) |
| **Survived**<br>**(n=161)** | 1,975.57<br>(1,646.94–<br>2,304.20) | 399.63<br>(360.33–<br>438.93) | 2,375.21<br>(2,020.75–<br>2,729.67) | 437.80<br>(365.06–<br>510.54) | 2,813.01<br>(2,421.53–<br>3,204.50) |
| ***Sequelae*** |  |  |  |  |  |
| **With sequelae**<br>**(n=64)** | 2,619.72<br>(1,968.95–<br>3,270.49) | 433.75<br>(377.56–<br>489.94) | 3,053.47<br>(2,362.78–<br>3,744.17) | 589.46<br>(439.27–<br>739.65) | 3,642.94<br>(2,878.64–<br>4,407.24) |
| **Without**<br>**sequelae**<br>**(n=100)** | 1,583.40<br>(1,260.49–<br>1,906.31) | 376.04<br>(322.99–<br>429.08) | 1,959.44<br>(1,600.84–<br>2,318.04) | 334.58<br>(272.16–<br>397.00) | 2,294.02<br>(1,908.45–<br>2,679.60) |

Abbreviations: CI, confidence interval; ND, not done.
<sup>a</sup>Data are presented as mean (95% CI).
<sup>b</sup>See Figure 1 for the estimation of costs of illness

## DISCUSSION

Despite the clear presence and clinical threat of paediatric encephalitis in Vietnam, limited information regarding its economic burden is available to support policymakers and physicians in prioritizing the resources for the improvement and execution of intervention strategies. Likewise, no data exist regarding the economic burden of paediatric encephalitis in Ho Chi Minh City, particularly in its major children’s hospital. Here, we describe the results of a prospective hospital-based study during 2020–2022, estimating the cost of illness of encephalitis in children.

The results show that paediatric encephalitis cases are associated with a substantial economic burden in Vietnam. Direct costs were the main cost driver, accounting for 84.6% of the total cost, particularly the direct medical cost during hospitalization. In terms of the direct non-medical costs the transportation cost during hospitalization and expenses related to childcare arrangements were the main drivers. Patients with NMDAR-antibody encephalitis, more severely ill patients, and patients with sequelae had higher total costs. This is likely due to the higher degree of severity, increased care requirements and ongoing sequelae seen in these patients.

Importantly, the total direct medical costs associated with non-ventilated and ventilated children suffering from encephalitis in our study were approximately 10 times and 22.0 times higher respectively than Vietnam’s annual average per capita health care spending in 2020 (US$166.2, 2020 prices). The direct medical costs during hospitalization paid by the patients themselves (not covered by government health insurance) was approximately six times higher than the average minimum wage per month. We also found that families incur notable direct non-medical costs (with a mean of US$398.56 per case). These direct non-medical costs are not covered by insurance, and therefore, the families have to pay for these expenses themselves. These numbers are concerning and highlight the risk of families incurring catastrophic health expenditures. In addition, the productivity cost during the hospitalization period was also much higher than the minimum earnings of the parent, posing an important loss to the family and society.

Our study only looked at the acute phase of encephalitis. However persistent neurologic effects are common following encephalitis, for example personality change, behavioural disorders, movement disorders, intellectual disability, learning disorders, blindness, paresis, and sleeping problems (29,30). Such disorders are likely to have long-lasting economic impacts to individuals, families and society. In addition, our study has other limitations. Firstly, it only investigated patients admitted to one hospital in Ho Chi Minh City, Vietnam. These cost estimates can not necessarily be generalised to every encephalitis case in Vietnam. For example, as a specialist centre, it is possible that the severity of cases and association with long-term sequelae may be higher in our sample. It is important to note that the current/standard approach to monetise productivity losses remains an area of debate, particularly regarding the valuation of unpaid work and whether or not to include the valuation of the productivity losses of children (31–33). The local COVID-19 regulations during the study period likely reduced the number of caregivers per patient and the associated costs associated with the caregivers. Finally, the cost data estimates calculated within this paper were based directly on the charges from the patients’ hospital bills and the costs related to the staff time were assumed to be captured by the charges for the different services. However, these charges do not necessarily reflect the economic value of the resources utilized for their care (34–36). In order to try and capture economic costs within this context, a cost-to-charge ratio is commonly applied to the charges (which is based on the ratio of the hospital’s (or department’s) expenses and what they charge) (37,38) but the data were not available to do this adjustment within this study.

## CONCLUSION

Our results show that the cost of illness of encephalitis in children is considerable and higher in more severe patients, patients with sequelae, and ventilated patients. Notably, we found that despite high health insurance coverage, patients and families still incur significant costs. Of note, many of the children in our study suffered with JEV, a vaccine-preventable disease, indicating the potential of preventative public health measures to impact and reduce these cost outcomes.

## Data Availability

All relevant data are within the manuscript and its Supporting Information files.

## Disclaimers

The funder had no role in study design, data collection and analysis, decision to publish, or preparation of the manuscript.

## Acknowledgments

We are indebted to the patients for their participations in this study.

